# Serious adverse events reported from the COVID-19 vaccines: A descriptive study based on WHO database

**DOI:** 10.1101/2021.03.23.21253433

**Authors:** Siddhartha Dutta, Rimple Jeet Kaur, Jaykaran Charan, Pankaj Bhardwaj, Praveen Sharma, Sneha Ambwani, Mainul Haque, Ankita Tandon, Jha Pallavi Abhayanand, Sanchi Sukhija, S V Suman, Sanjeev Misra

## Abstract

**Background:** In the light of current pandemic, the emergency approval of few COVID-19 vaccines seems to provide a ray of hope. However, their approval is solely based on limited data available from the clinical trials in a short period of time; thereby imposing a necessity to study the adverse events(AEs) associated with their use. This study therefore aims to assess the Serious Adverse Events (SAEs) associated with various COVID-19 vaccines reported in the WHO database (VigiBase®).

**Methods:** The data from VigiBase® was analyzed to assess the reported SAEs linked to various COVID-19 vaccines. The duplicates in the data were removed and were analyzed on the basis of age, gender, and seriousness of adverse events at the System Organ Classification (SOC) level and the individual Preferred Term (PT) level.

**Results:** A total 103954 adverse events reported from 32044 subjects were taken for analysis. Of 32044 subjects, majority were females (80%). Also, a total of 28799 (27.7%) SAEs were reported from the 8007 individuals. Most of the SAEs were observed in Europe (83%) amongst females (79.4%) between 18 to 64 years (80.74%) of age. Majority of SAEs (74%) were reported for BNT162b2 (Pfizer) vaccine. On system wise classification, general disorders (30%) were the commonest followed by nervous system (19.1%) and musculoskeletal (11.2%) disorders. In individual category, headache (8.1%) was the commonest, followed by pyrexia (7%) and fatigue (5.1%). The number of SAEs were reported with various vaccines were comparatively lesser as compared to the non-serious ones and incidence of death was low with all the vaccines candidates. Elderly (> 65 years) people reported more serious AEFIs as compared to other age groups.

**Conclusion:** The reported SAEs from the COVID–19 vaccines were in line with the data published in clinical trials. To link these SAEs to vaccines will need causality analysis and review of individual reports.

## Introduction

The ongoing COVID-19 pandemic, caused by severe acute respiratory syndrome coronavirus 2 (SARS-CoV-2) has enveloped the entire globe with serious impositions on almost every phase of human life.^1, 2^ The World Health Organization (WHO) reported approximately 99,864,391 cases of COVID-19 alongside surmounting to nearly 2,149,700 deaths by January 27^th^, 2021.^3^ The rapid upsurge in active COVID-19 cases produced a major health care crisis due to lack of alertness to confront a sudden pandemic especially in developing nations.^4^ However, in accordance with the present situation, there has been a downward trend in the number of COVID-19 cases as reported by WHO.^5^

At the onset, the disease had multifaceted spectrum with no effective medication, hence hit and trial strategy with older drugs was implanted in order to find an answer to this morbid disease.^6^ However a lot of therapies were being tried and are still under clinical trials to prove their efficacy yet infection control measures, sanitation, symptomatic and supportive therapy has been the cornerstone of effective clinical management of the COVID-19.^6,7^ Later, Food and Drug Administration (FDA) approved Remdesivir for the treatment of COVID-19.^8^

As there was no definitive therapy in conjunction with a tremendous rise in the number of cases, an effective vaccine vestiges the only answer in building immunity in order to bring a halt to further progression of the disease.^9^ As of Jan 26, 2021, WHO in its “COVID-19 - Landscape of novel coronavirus candidate vaccine development worldwide” document reported a total of 236 vaccine candidates under development and of which 63 were in clinical trial phase and 173 were under preclinical development.^10^ The vaccines that are in clinical development are mostly protein subunit vaccine, Viral Vector (non-replicating), DNA, Inactivated Virus and RNA vaccines.^10^ With most of the trials underway and few clearing phase 3 studies, the pharmaceutical industries applied for the Emergency Use Authorization (EUA) of few vaccines. As of January 29, 2021 there were a total of nine vaccines which have been approved around the world namely Comirnaty (BNT162b2), Moderna COVID-19 Vaccine (mRNA-1273), CoronaVac, COVID-19 Vaccine AstraZeneca (AZD1222), vaccine from Sinopharm and the Wuhan Institute of Virology, Sputnik V, BBIBP-CorV, EpiVacCorona and Covaxin.^11^ The FDA on Dec 11, 2020 gave EUA to the Pfizer-BioNTech vaccine for COVID-19 to be distributed in the United States of America for individuals having age 16 years of and older.^12^ Later by Dec 18, 2020 FDA also approved Moderna vaccine for COVID-19 for use for in individuals 18 years of age and older.^13^ However, FDA emphasized that EUA is only based on limited efficacy safety data and it is not full approval of the vaccine.^12, 13^ In India, two vaccines namely Covishield from Serum Institute of India and Covaxin from Bharat Biotech received Restricted Emergency Approval for prevention of COVID-19.^14^ Although these vaccines have been granted approval for emergency use, their long term efficacy is yet to be established.

It is extremely critical to monitor the vaccine safety and/or SAEs using Pharmacovigilance which is defined as “the science and activities related to the detection, assessment, understanding and prevention of adverse effects or any other drug related problem”.^15^ Pharmacovigilance is an essential component while using any medical or paramedical modalities for patient care and safety as monitoring adverse effects of drugs, vaccines and other interventions would help identify the safer drugs, preventing patients from unnecessary harms and reducing the hospitalization and treatment cost hence assuring rational use of medications and interventions.^16^ For strict pharmacovigilance WHO authorizes and maintains global database of AEFIs through VigiBase® which maintains the global safety data of various therapeutic interventions as Individual Case Safety Reports (ICSRs).^17^ It came into existence in 1968 and consists of over 20 million ICSR from over 130 countries. ICSRs are also known as the spontaneous or voluntary reports which are generated in the post marketing phase of the drug. Each ICSR contains the information regarding patient’s demographics; drugs; adverse events and administrative information.^17, 18^

This descriptive analysis is an extension to the studies conducted using same database on drugs used in COVID-19 therapeutics including Favipiravir and Remdesivir.^19, 20^ This descriptive study aims to identify and describe various SAEs reported for the COVID – 19 vaccines through WHO database thereby facilitating identification of safer vaccines, preventing patients from unnecessary tribulations and reducing the hospitalization and treatment costs in order to assure rational vaccination regimens and strategies.

## Methodology

### Data Source

Data for this study was obtained from the VigiBase®, which is a database maintained by WHO Uppsala monitoring center (UMC), Uppsala, Sweden. All vaccine safety data reported from 15^th^ December 2020 to 24^th^ January 2021 were obtained in excel format from the UMC. Data were cleaned from duplicates and irrelevant entries by the first and second author (SD, RK) and any discrepancy for removal or retention of data for analysis was resolved by discussion and consensus in the presence of corresponding author (JC). This database has all the data reported in the form of adverse drug events associated with COVID-19 vaccines using Individual Case Safety Reports (ICSRs).^17^ The detailed information regarding patient’s demographics (age, gender, country, medical history); drugs (indication of use, route of administration, start and end date); adverse events (date of onset, seriousness, outcome, dechallenge and rechallenge outcomes. and causality) and administrative information (type and source of report) was recorded.^17, 21^

### Data interpretation and analysis

Each report in VigiBase® represents an individual Adverse Events (AEs) and there could be more than one report for a single individual thus the number of reported AEs were more than the number of individuals who actually had adverse event. Hence, the data was cleaned manually to remove the duplicates in the form of same adverse events reported for same individual in different terminologies.

All the adverse drug reactions in the ICSR are automatically coded as per MedDRA (Medical Dictionary for Regulatory Activities) and WHO-ART terminology.^17,21^ MedDRA is the hierarchical terminology that is composed of five levels: Lowest Level Terms (LLTs), Preferred Term (PTs), High Level Group Terms (HLGTs), High–Level Terms (HLT) and SOCs (System Organ Classification).^22, 23^

In the present study, the SOC and PT categories of AEs were only employed for the analysis. Here PT refers to clinical condition in the form of symptom, sign, diagnosis, investigation, medical, social or family history and characteristic surgical or medical procedures.^23^ Each PT is linked to specific SOC which is grouping by manifestation site (eg. Cardiovascular disorder), aetiology (infections and infestations) and purpose (surgical and medical procedures).^24^ The age, gender and severity of all the adverse events were compared with the SOC and PT. The seriousness of the adverse event was decided as per the ICH E2B criteria which identifies SAEs as those leading to either life threatening event, hospitalization, disability, congenital abnormality or death.^25^

### Ethical Approval

This study had no direct interaction with human participants and was based on the WHO’s database (VigiBase®) hence, the ethical approval was not required.

### Statistical Analysis

Descriptive statistics were reported in the form of frequency and percentages. Cross tabulation function of Statistical Package for Social Science version 17 was used for the analysis.

## Results

A total 103954 AEs were reported till 24^th^ Jan 2021 from 32044 subjects (Average 3.24 AEs per person). Out of the total subjects, 5731 (17.9%) were males and 25652 (80 %) were females. A total of 28799 (27.7%) AEs from 8007 individuals were categorized as Serious Adverse Events (SAEs) **(Figure 1)**. Majority of SAEs were reported in females and between the age group of 18 to 64 years. Around 83% of the SAEs were reported from European countries(N=23987) followed by Americas(N=4795) and Asia(N=17) **(Figure 2)**. In almost 74% of cases the BNT162b2 (Pfizer) vaccine was used. Around 1% of SAEs were fatal **(Table 1)**.

**Figure 1.**
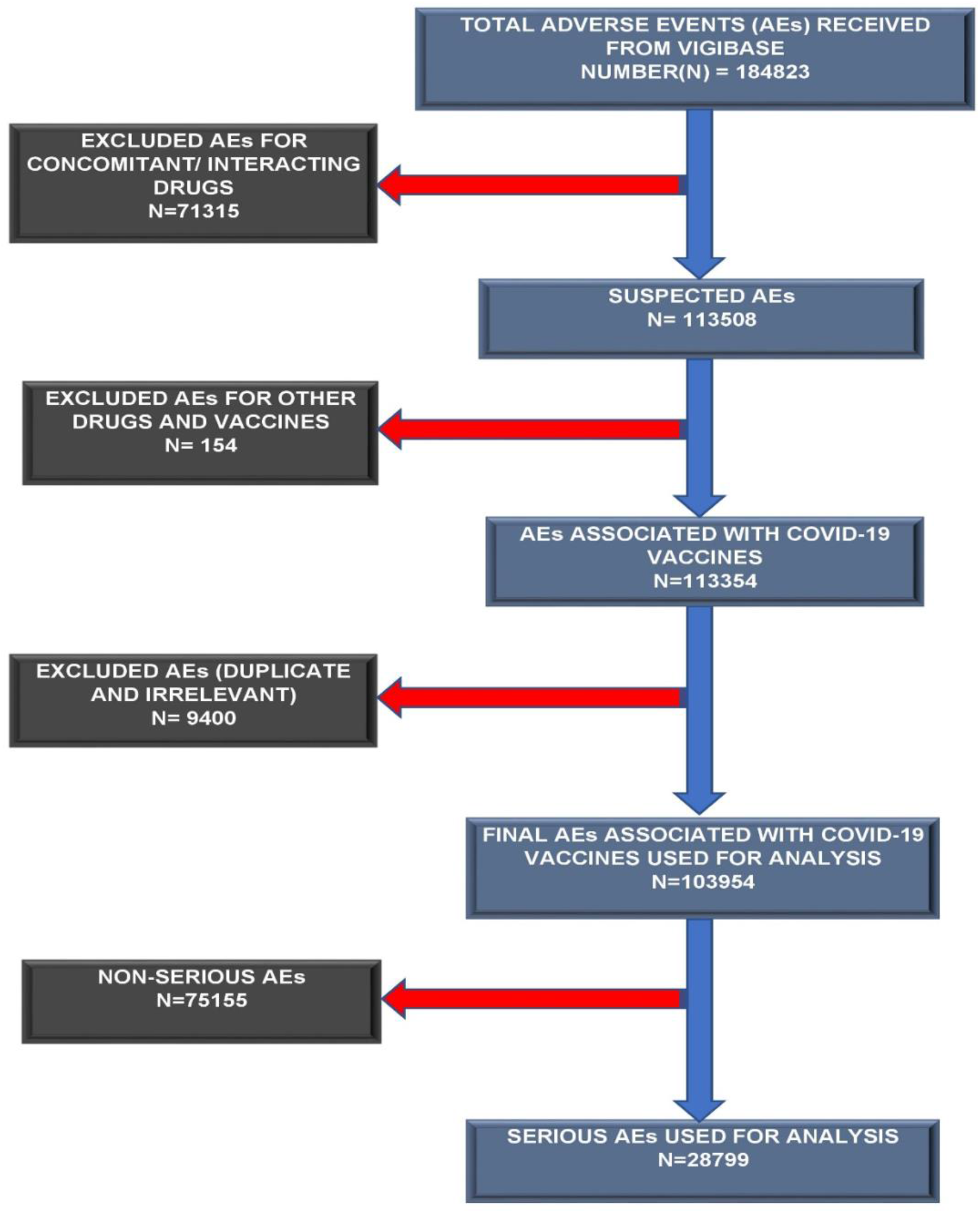
Schematic Diagram of assessment of Adverse Events associated with COVID-19 vaccines in VigiBase database

**Figure 2.**
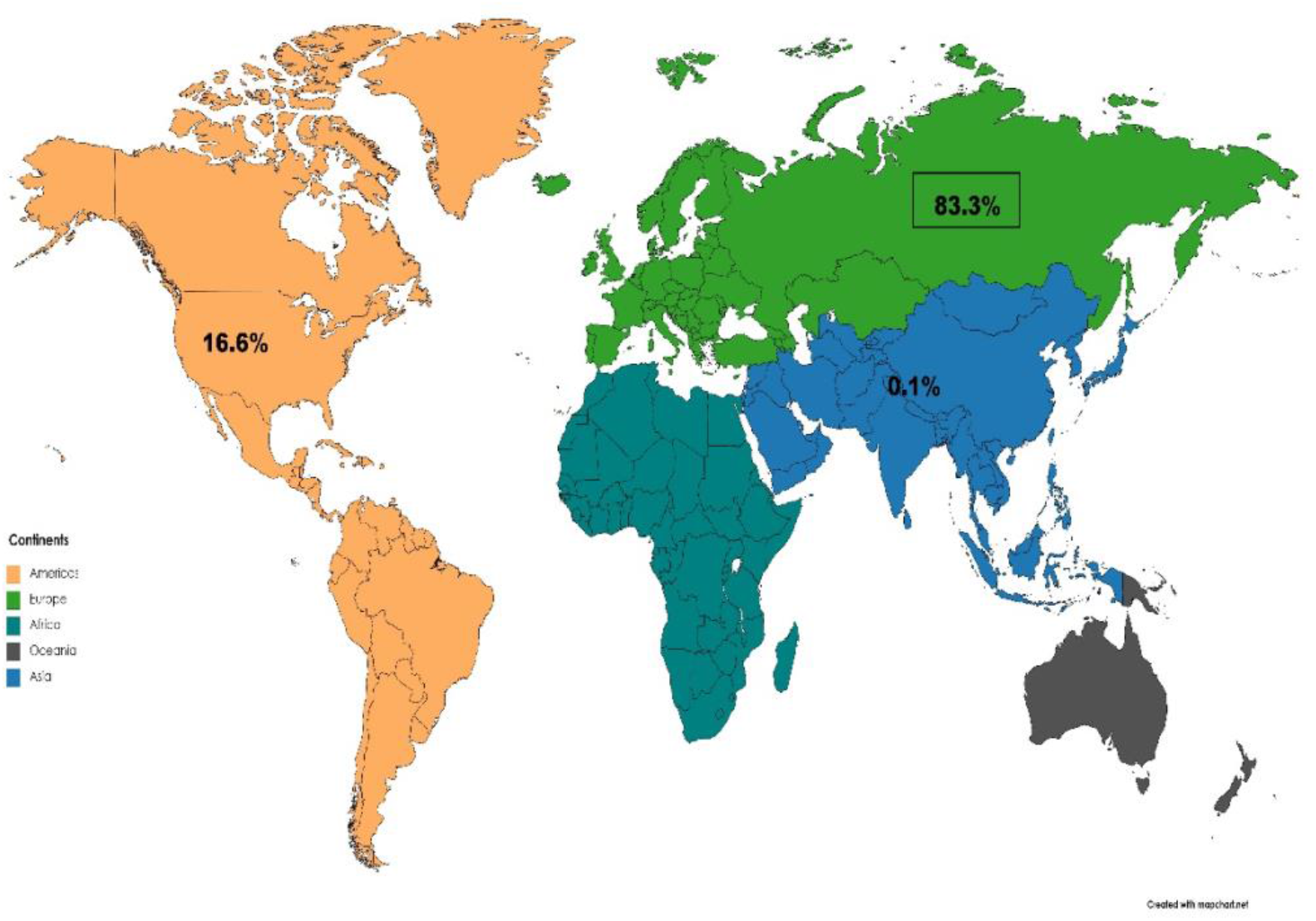
Distribution of Serious Adverse Events reported in VigiBase associated with COVID-19 vaccines across continents

**Table 1:**
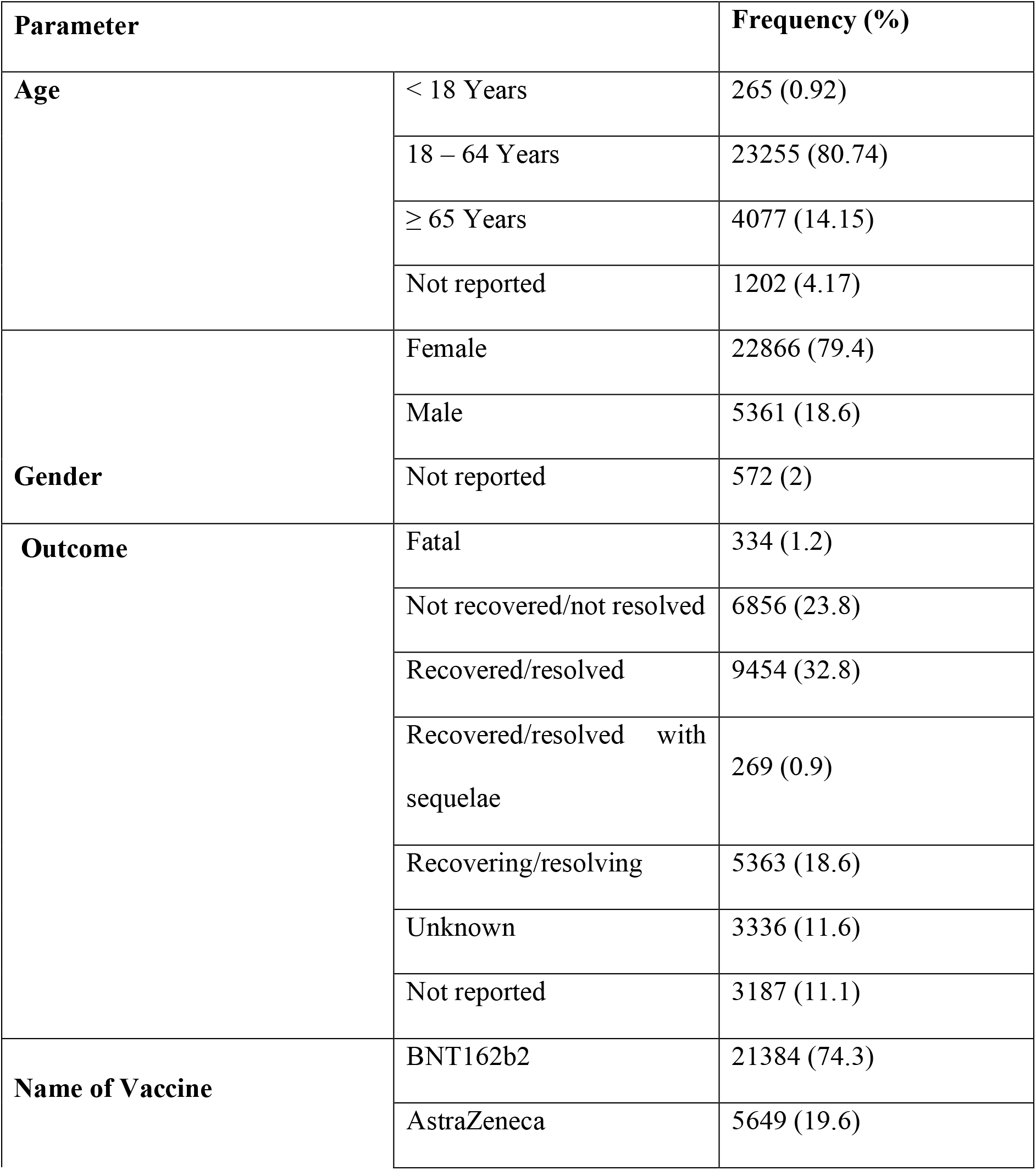
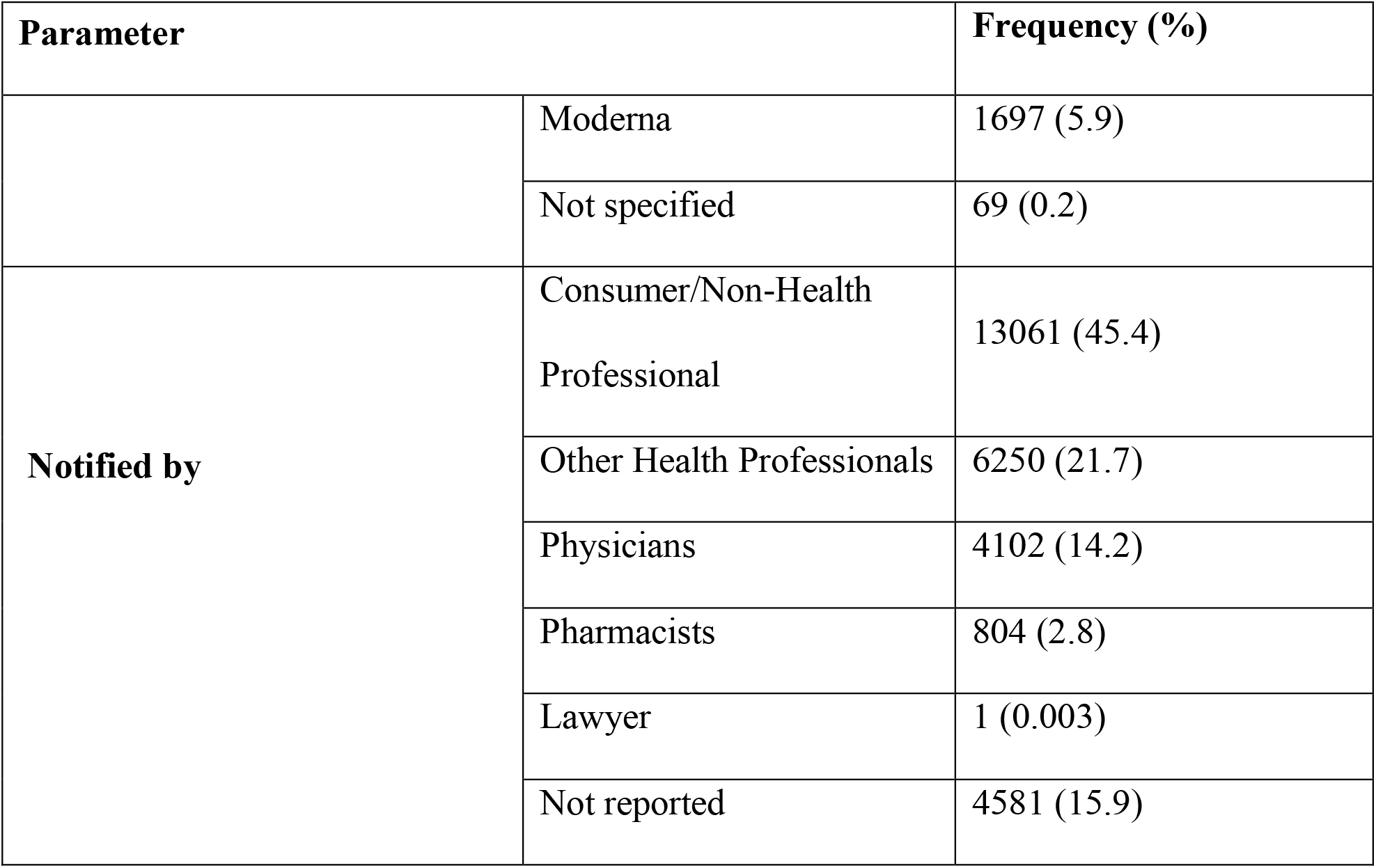
Characteristics of SAEs (28799 SAEs reported from 8007 Individuals) reported for COVID – 19 vaccines in WHO database (n = 28799)

| **Parameter** |  | **Frequency (%)** |
| --- | --- | --- |
| **Age** | < 18 Years | 265 (0.92) |
|  | 18 – 64 Years | 23255 (80.74) |
|  | ≥ 65 Years | 4077 (14.15) |
|  | Not reported | 1202 (4.17) |
| **Gender** | Female | 22866 (79.4) |
|  | Male | 5361 (18.6) |
|  | Not reported | 572 (2) |
| **Outcome** | Fatal | 334 (1.2) |
|  | Not recovered/not resolved | 6856 (23.8) |
|  | Recovered/resolved | 9454 (32.8) |
|  | Recovered/resolved with sequelae | 269 (0.9) |
|  | Recovering/resolving | 5363 (18.6) |
|  | Unknown | 3336 (11.6) |
|  | Not reported | 3187 (11.1) |
| **Name of Vaccine** | BNT162b2 | 21384 (74.3) |
|  | AstraZeneca | 5649 (19.6) |

| Parameter |  | Frequency (%) |
| --- | --- | --- |
|  | Moderna | 1697 (5.9) |
|  | Not specified | 69 (0.2) |
| Notified by | Consumer/Non-Health Professional | 13061 (45.4) |
|  | Other Health Professionals | 6250 (21.7) |
|  | Physicians | 4102 (14.2) |
|  | Pharmacists | 804 (2.8) |
|  | Lawyer | 1 (0.003) |
|  | Not reported | 4581 (15.9) |

Majority of the SAEs were reported from the broad category “general disorders and administration site conditions” (30%), followed by “Nervous system disorders” (19.1%), “Musculoskeletal and connective tissue disorders” (11.2%) and “Gastrointestinal disorders” (10.7%) **(Table 2)**.

**Table 2:** Distribution of SAEs reported from the COVID – 19 vaccines as per the broad system-based classification (N=28779).

| **Adverse Events Broad Heading Categories** | **Frequency (%)** |
| --- | --- |
| Blood and lymphatic system disorders | 355 (1.23) |
| Cardiac disorders | 710 (2.47) |
| Congenital, familial and genetic disorders | 2 (0.01) |
| Ear and labyrinth disorders | 191 (0.66) |
| Eye disorders | 365 (1.27) |
| Gastrointestinal disorders | 3103 (10.77) |
| General disorders and administration site conditions | 8640 (30.0) |
| Hepatobiliary disorders | 12 (0.04) |
| Immune system disorders | 363 (1.26) |
| Infections and infestations | 565 (1.96) |
| Injury, poisoning and procedural complications | 82 (0.28) |
| Investigations | 1006 (3.49) |
| Metabolism and nutrition disorders | 234 (0.81) |
| Musculoskeletal and connective tissue disorders | 3225 (11.20) |
| Neoplasms benign, malignant and unspecified (incl cysts and polyps) | 3 (0.01) |
| Nervous system disorders | 5502 (19.10) |
| Pregnancy, puerperium and perinatal conditions | 17 (0.06) |
| Product issues | 1 (0.003) |
| Psychiatric disorders | 470 (1.63) |
| Renal and urinary disorders | 83 (0.29) |
| Reproductive system and breast disorders | 59 (0.20) |
| Respiratory, thoracic and mediastinal disorders | 1709 (5.93) |
| Skin and subcutaneous tissue disorders | 1500 (5.21) |
| Social circumstances | 22 (0.08) |
| Surgical and medical procedures | 20 (0.07) |
| Vascular disorders | 560 (1.94) |

Upon analyzing the Preferred Terms (PTs) in the broad categories, 28799 SAEs were reported. Headache (8.11%) of various types were most common SAEs followed by pyrexia (7.09%), fatigue (5.18%), nausea (4.4%), chills (4.2%) and myalgia (3.9%). Pain (1.93 %), pain in extremity (1.98 %) and vaccination site /injection site/administration site pain (1.88) accounted for another major portion of SAEs **(Table 3)**.

**Table 3:**
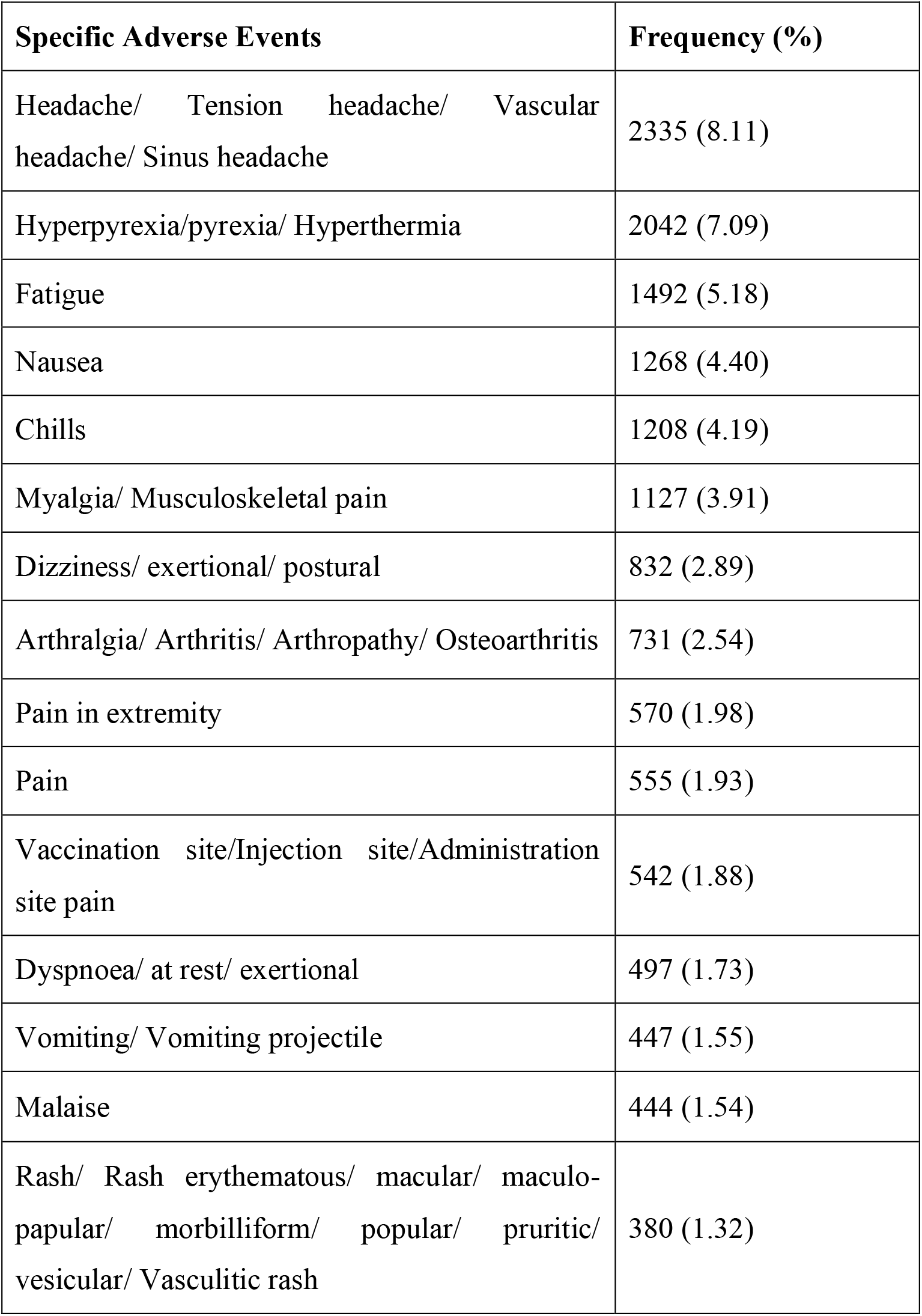
Common individual Adverse Events reported from the COVID – 19 vaccines.

| **Specific Adverse Events** | **Frequency (%)** |
| --- | --- |
| Headache/ Tension headache/ Vascular headache/ Sinus headache | 2335 (8.11) |
| Hyperpyrexia/pyrexia/ Hyperthermia | 2042 (7.09) |
| Fatigue | 1492 (5.18) |
| Nausea | 1268 (4.40) |
| Chills | 1208 (4.19) |
| Myalgia/ Musculoskeletal pain | 1127 (3.91) |
| Dizziness/ exertional/ postural | 832 (2.89) |
| Arthralgia/ Arthritis/ Arthropathy/ Osteoarthritis | 731 (2.54) |
| Pain in extremity | 570 (1.98) |
| Pain | 555 (1.93) |
| Vaccination site/Injection site/Administration site pain | 542 (1.88) |
| Dyspnoea/ at rest/ exertional | 497 (1.73) |
| Vomiting/ Vomiting projectile | 447 (1.55) |
| Malaise | 444 (1.54) |
| Rash/ Rash erythematous/ macular/ maculopapular/ morbilliform/ popular/ pruritic/ vesicular/ Vasculitic rash | 380 (1.32) |

**Table 4** describes the distribution of serious and non-serious AEs between type of vaccines, gender and age groups. The comparison of serious vs. non serious AEs with various vaccine candidates shows the probability of serious AEs are comparatively low as compared to the non-serious AEs. Age group > 65 years had more serious AEs as compared to other age groups. There was equal distribution of serious and non-serious AEs between males and females. There were a total 424 deaths. Distribution of these deaths as per the vaccines, gender and age group have been mentioned in the **Table 5**.

**Table 4:** Distribution of serious and non-serious Adverse Events as per the vaccine, gender and age groups.

|  |  | **Serious** | **Non-Serious** | **Total** |
| --- | --- | --- | --- | --- |
| **Vaccine Name**<br>**(N = 103954)** | BNT162b2(Pfizer) | 21384<br>(25.23) | 63357<br>(74.77) | 84741 |
|  | AstraZeneca | 5649 (45.15) | 6861 (54.84) | 12510 |
|  | Moderna | 1697 (26.73) | 4650 (73.27) | 6347 |
|  | Covid-19 vaccine (Vero cell),<br>inactivated | 9 (28.12) | 23 (71.87) | 32 |
|  | Covaxin | 0 | 2 (100) | 2 |
|  | No vaccine name mentioned | 60 (18.7) | 261 (81.30) | 321 |
| **Gender**<br>**(N = 103954)** | Male | 5361 (29.30) | 12935<br>(70.70) | 18296 |
|  | Female | 22866<br>(27.41) | 60551<br>(72.59) | 83417 |
|  | Gender not reported | 572 (25.52) | 1669 (74.48) | 2241 |
| **Age Group**<br>**(N = 103954)** | < 18 Years | 265 (31.29) | 582 (68.71) | 847 |
|  | 18 – 64 Years | 23255<br>(25.98) | 66250<br>(74.02) | 89505 |
|  | >65 Years | 4077 (53.32) | 3570 (46.68) | 7647 |
|  | Unknown | 1202 (20.18) | 4753 (79.82) | 5955 |

Values in parenthesis are percentages. N= total adverse events

**Table 5:** Distribution of events of death as per the vaccine, age and gender (N = 103954).

|  |  | **Death (%)** |
| --- | --- | --- |
| **Vaccine Name** | BNT162b2 (N=84741) | 337 (0.40) |
|  | AstraZeneca (N = 12510) | 9 (0.07) |
|  | Moderna (N = 6347) | 78 (1.23) |
| **Gender** | Male (N = 18296) | 205 (1.12) |
|  | Female (N = 83417) | 213 (0.25) |
|  | Gender not reported (N=2241) | 6 (0.27) |
| **Age Group** | < 18 Years (N=847) | 3 (0.35) |
|  | 18 – 64 Years (N=89505) | 43 (0.05) |
|  | >65 Years (N = 7647) | 342 (4.47) |
|  | Unknown (N = 5955) | 36 (0.6) |

## Discussion

The present study was conducted to assess the SAEs reported in the global pharmacovigilance database (VigiBase®) associated with various COVID-19 vaccines like BNT162b2(Pfizer), AZD1222/ChAdOx1 nCoV-19(AstraZeneca), Moderna etc. that are currently being used across the world for vaccination. Approximately one third of the total AEs reported were serious in nature. The majority of the AEs reported were from the female subjects and from age group 18 to 64 years. A major chunk of AEs was reported from Europe and in majority of the cases BNT162b2(Pfizer) vaccine was used. Most commonly the AEs reported were classified under general disorders and administration side conditions with headache, fever and fatigue as the commonest AEs observed.

In the current analysis, the ratio of serious and non-serious AEs was similar amongst males and females but overall, more numbers were reported from females. Centres for Disease Control and Prevention(CDC) in their Morbidity and Mortality Weekly Report (MMWR) reported that after first dose of Pfizer-BioNTech COVID-19 Vaccine almost 90 % of the anaphylactic reactions and non-anaphylactic allergic reactions were observed in females among which 81% of them with anaphylaxis and 67% with non-anaphylactic allergic reactions had a history of allergic reactions.^26,27^ However in the above report, about 62% of the Pfizer-BioNTech COVID-19 vaccines were received by females which can be a reason for preponderance of allergic reactions in females.^26^ Similar report by CDC on the Moderna COVID-19 Vaccine reported that about 61% of the vaccine recipients were females and almost 100% of the anaphylactic reaction and 91% of non-anaphylactic allergic reactions were observed in females amongst which 90% of them with anaphylaxis and 60% with non-anaphylactic allergic reactions had a history of allergic reactions.^28^

The proportion of female population in United States was 50.51% and in Europe it was 51.12 % in 2019.^29, 30^ WHO in its ‘Health Workforce Working paper 1’ compared the percentage of male and female health care professionals(HCPs) in Europe and America in 2019 and the percentage of female physicians in USA was 46% but was 53% in Europe whereas the percentage of nursing workforce was specifically dominated by females which was 86% in USA and 53% in Europe.^31^ So the more AEs reported in females could be attributed to a greater number of females as health workers. The During the early roll out of the vaccine, it was preferably given to the Health Care Professionals (HCPs) and the above data shows a female predominance in the HCPs hence more female HCPs might have received the vaccine as compared to the male ones which could be a reason for the more number AEs reported by the females. However, the difference in the reporting is too skewed to be justified only on the basis of this reason.

Study conducted by Harris T et al on assessing the gender-specific differences in AEs reporting with various vaccines in Ontario during 2012-2015 reported that the most of the AEs reported (66.2%) were associated with females.^32^ However, there was a more even distribution observed while analysing the SAEs of either gender (57.5% female, RRR 1.3).^32^ Literature reveals that previous studies done by Huang et al (63%) on MF59®-adjuvanted H5N1 influenza vaccine and Halsey et al on H1N1 vaccines reported female predominance in the AEs associated with the respective vaccinations.^33, 34^ A review conducted by Fink et al on impact of gender and response to vaccines in elderly reported that the AEs with females as compared to males were consistently higher with response to various vaccines like influenza, pneumococcal, herpes zoster, tetanus and pertussis.^35-41^ The reactions observed by the either gender were similar but female vaccine recipients reported more local reactions, like injection site pain, redness, and swelling, as well as some of the systemic reactions like joint pain, myalgia, headache, back pain, abdominal pain, fever, chills, and hypersensitivity reactions.^35, 41^ Few probable explanations in favour of females with higher number of AEs can be due to increased humoral and cell-mediated immune reactions to antigens, vaccine, and infections as compared to males.^42, 43^

The SAEs with the Pfizer-BioNTech COVID-19 Vaccine from their clinical trial experience in the 16 to 55 years of age were reported by 0.4% of the recipients and 0.8% of the participants with more than 56 years of age and older.^44^ In the present analysis, the SAEs constituted of 25.23% of the total AEs reported in the VigiBase and deaths were observed in 0.40% of total SAEs associated with Pfizer-BioNTech vaccine. Whereas, as per the data reported from clinical trials, death was reported in two (0.01%) vaccine recipients and both of them were above 55 years of age.^44, 45^ The proportion of non-fatal SAEs was 0.6% with the vaccine and most common AEs reported were appendicitis (0.04%), acute myocardial infarction (0.02%), and cerebrovascular accident (0.02%).^44^

As per the clinical trial experience of Moderna COVID-19 Vaccine, the proportion of vaccine recipients who developed at least one AEs was 1%. In our analysis the SAEs constituted of 26.73% of the total AEs reported in the VigiBase and death was observed in 1.23% of total SAEs associated with Moderna vaccine. As per the data reported to FDA there were six deaths reported after vaccination and most of them were above 70 years of age and associated co-morbid conditions. In the vaccine recipient group, the commonest SAEs reported were myocardial infarction (0.03%), cholecystitis (0.02%), and nephrolithiasis (0.02%). As per the opinion of FDA, three SAEs were also considered to be likely caused by the vaccines which were one case of intractable nausea/vomiting and two of facial swelling.^46, 47^ The discrepancy in the proportion of SAEs between our study and the one reported to the FDA could be due to the fact that we calculated the proportion out of total adverse events reported and not from the total patients vaccinated.

The AEs from the COVID-19 vaccine from AstraZeneca as reported to Medicines & Healthcare products Regulatory Agency (MHRA) were not classified as serious or non-serious but reported general disorders and administration site conditions like injection site reaction/pain, fatigue, headache and nausea to be commonest SAEs ^48^ A recent study published based on an interim analysis of four clinical trials conducted in Brazil, South Africa, and UK has also reported that 79(0.7%) of whom received ChAdOx1 nCoV-19 suffered gastrointestinal disorders, injury, poisoning and procedural complications, infections/infestations and nervous system disorders.^49^ In our analysis more serious adverse events were reported in older age group in comparison to the younger age groups, this warrant a cautious approach when administrating vaccine to the old age group people in the form of longer follow up and watchful vigilance.

The data analyzed in this study has been adapted from VigiBase which is a WHO global database for individual case safety reports. The data is collected from several sources and the probability that the suspected adverse effect is drug-related is not the same in all cases. In the absence of proper reporting of other parameters and technical problems in causality assessment, it is not appropriate to attribute all of these events to vaccine hence in this paper we used the term adverse events and not the adverse drug reaction which is more definitely linked to the drug. The data presented in this study does not represent the opinion of the Uppsala Monitoring Centre or the WHO.

## Conclusion

The present study that the pattern of AEs reported in the database was in sync with the reactogenicity of the vaccines. However, there is an urgent need of systematic analysis regarding different AEs reported in this study for the purpose of measuring causalities through proper review of reports and by generating data in primary studies.

## Data Availability

All the data related to the study is available with the corresponding author

## Author Contributions

All authors made a significant contribution to the work reported, whether that is in the conception, study design, execution, acquisition of data, analysis, and interpretation, or in all these areas; took part in drafting, revising, or critically reviewing the article; gave final approval of the version to be published; have agreed on the journal to which the article has been submitted; and agree to be accountable for all aspects of the work.

## Acknowledgment

We want to acknowledge ‘mapchart.net’, a free service used for preparation of a map diagram for this study

